# Seizure prediction in 1117 neonates leveraging EMR-embedded standardized EEG reporting

**DOI:** 10.1101/2022.06.03.22275975

**Authors:** Jillian L. McKee, Michael C. Kaufman, Alexander K. Gonzalez, Mark P. Fitzgerald, Shavonne L. Massey, France Fung, Sudha K. Kessler, Stephanie Witzman, Nicholas S. Abend, Ingo Helbig

## Abstract

**Background:** Accurate prediction of seizures can help direct resource-intense continuous EEG (CEEG) monitoring to high-risk neonates. We aimed to use data extracted from standardized EEG reports to generate seizure prediction models for vulnerable neonates.

**Methods:** In 2018, we implemented a novel CEEG reporting system in the electronic medical record (EMR) that incorporated standardized terminology. We developed seizure prediction models using logistic regression, decision tree, and random forest models for neonates and specifically, neonates with hypoxic-ischemic encephalopathy (HIE), using EEG features on day 1 to predict future seizures.

**Findings:** We evaluated 1117 neonates, including 150 neonates with HIE, with CEEG data reported using standardized templates. Implementation of a consistent EEG reporting system, which documents discrete and standardized EEG variables, resulted in >95% reporting of key EEG features. Several EEG features were highly correlated, and patients could be clustered based on specific features. However, no simple combination of features adequately predicted seizure risk. We therefore applied computational models to complement clinical identification of high-risk neonates. Random forest models incorporating background features performed with classification accuracies of up to 90% for all neonates and 97% for neonates with HIE, and recall (sensitivity) of up to 97% for all neonates and >99% for neonates with HIE.

**Interpretation:** Using data extracted from the standardized EEG report on the first day of CEEG, we predict the presence or absence of neonatal seizures on subsequent days with classification performances of >90%. This information, incorporated into routine care, can guide decisions about the necessity of continuing CEEG beyond the first day and thereby improve the allocation of limited CEEG resources. Additionally, this analysis illustrates the benefits of standardized clinical data collection which can drive learning health system approaches to personalized CEEG utilization.

**Funding:** Children’s Hospital of Philadelphia, The Hartwell Foundation, NINDS, Wolfson Foundation

**Research in context:** *Evidence before this study:* We searched the literature on EEG-based seizure prediction among neonates in PubMed from January 1, 1946, to June 1, 2022, using combinations of the keywords “seizure,” “prediction,” “EEG,” “neonatal,” and “hypoxic-ischemic encephalopathy.” We used no language restrictions. Prior studies relied on manual review of EEG reports to forecast seizures in neonates using regression models. These studies were limited in sample size as they required manual review of reports and manual data entry. No studies were identified using automated collection of EEG data from routine care, and none used machine learning-based modeling techniques.

*Added value of this study:* We built seizure prediction models based on standardized EEG features reported in the EMR which could predict seizures in neonates, and particularly those with HIE, with greater than 90% accuracy. Furthermore, these models could be tuned to not miss seizures, performing with recall (sensitivity) of up to 97% in the overall neonatal cohort and >99% among neonates with HIE, while still maintaining precision (positive predictive value) of up to 92% and 97%, respectively. Previous studies have built seizure-prediction models using EEG data, but most have used features derived from manual scoring of EEG tracings or computational analysis of the raw EEG recordings. While these studies are informative, they are not easily scalable for incorporation into routine clinical practice. To our knowledge, this is the first study reporting a seizure-prediction model based on standardized reports already documented in the EMR that can be used for clinical decision support to improve care for critically ill neonates. Prediction models developed in our study are available at http://neopredict.helbiglab.io.

*Implications of all the available evidence:* Continuous EEG monitoring is currently the standard of care for critically ill children at increased risk of seizures. While effective for seizure detection, long-term monitoring is resource-intensive and can have physical and psychosocial consequences, such as skin breakdown and reduced bonding. Accurately predicting which neonates are likely to seize after an initial shorter period of monitoring would help allocate resources towards neonates at highest risk of seizures and avoid unnecessary use of limited EEG monitoring resources in neonates at low risk of seizures. Furthermore, the ability to directly extract these predictors from the EMR will allow for automated predictions and dashboard development for use at scale and in real-time in clinical care.

## Introduction

Neonatal seizures are common and contribute significantly to morbidity and mortality.^1^ Neonates with hypoxic-ischemic encephalopathy (HIE) have a high incidence of seizures (∼30%),^2–4^ which are mostly EEG-only, and thus would not be identified by clinical observation alone. Additionally, among neonates with HIE, seizures have been associated with an increased risk for subsequent neurobehavioral problems and epilepsy.^2,4–7^ Due to the high risk of seizures in this population, guidelines recommend that neonates with HIE undergo continuous EEG monitoring (CEEG) liberally during the therapeutic hypothermia and return to normothermia periods, which often last 4-5 days.^8,9^ However, this practice is resource-intensive and not feasible for all neonates who might benefit,^10^ as many neonates receive care in neonatal intensive care units without CEEG capability and approaches to remote CEEG are new and not yet widely available.^11^ Furthermore, CEEG is not entirely benign, as long-term electrode placement can cause skin breakdown^12^ and the necessary wires and head-wrap can interfere with maternal-infant bonding and feeding.^13^ Therefore, it would be helpful to predict each individual’s risk of seizure to aid in the allocation of resources and minimize unnecessary medical procedures, shifting focus to neonates at highest risk.

Prior studies have shown that the prediction of neonatal seizures is complex, and clinical and EEG data generally do not predict seizures well. Clinical features alone are not predictive of seizures in neonates^3,14^ and EEG studies have found that while normal backgrounds correctly predict the absence of seizures,^15^ an abnormal background does not accurately predict the presence of seizures.^3,15–17^ Seizure prediction models have been developed based on EEG features determined by manual review of EEG segments,^18–21^ review of EEG reports,^16,17^ or by direct computational analysis of EEG tracings.^22^ These models have been limited by the number of individuals or the short windows of review,^23^ and only a few have been performed in neonates.^16,17,19–21^ Due to the need for manual chart or EEG review, these existing models cannot easily be incorporated into routine clinical care.

We have implemented a standardized reporting template for all clinical EEG reports which is derived from terminology published by the American Clinical Neurophysiology Society (ACNS),^24^ and have previously demonstrated that this reporting system is acceptable to electroencephalographers.^25^ Here, we aimed to determine whether this standardized template leads to complete reporting of the recommended terminology, and subsequently aimed to develop neonatal seizure prediction models based on data extracted from these reports to optimize CEEG utilization among neonates, including the subset with HIE.

## Methods

### Clinical Management

Patients were managed by neonatology and neurocritical care consultation services, and CEEGs were interpreted by pediatric electroencephalographers knowledgeable regarding the ACNS standardized terminology.^24^ Prophylactic anti-seizure medications (ASMs) were not administered but some neonates were receiving ASMs if they had experienced clinically-evident seizures prior to CEEG initiation. CEEG was performed following an institutional CEEG pathway consistent with ACNS recommendations^8^ and generally lasted 2 days among most neonates, 4-5 days among neonates with HIE, and at least 24 hours after the last seizure among neonates with seizures. All neonates who experienced electrographic seizures while being monitored on CEEG were treated with appropriate ASMs.

### EEG features were acquired using CDEs extracted from the EMR

In January 2018, our center implemented a novel EEG reporting system based on common data elements (CDEs) in the electronic medical record (EMR) that incorporated standardized terminology from the ACNS.^24^ The neonatal template included embedded descriptions of continuity, variability, reactivity, voltage, graphoelements, epileptiform transients, seizures, and an overall impression (**Figure S1**). Details regarding the reporting system have been published previously.^25^ These CDEs, along with demographic features and diagnosis codes, were extracted from the EMR for analysis.

### Patients were selected based on demographic factors, diagnoses codes, and EEG study type

All neonatal CEEG data were extracted from the EMR using age at the time of EEG and the EEG study type CDEs. Individuals were included in this cohort if they underwent CEEG during the first 30 days of life. Neonates with HIE were selected from the EMR using the ICD-10 diagnosis codes for “hypoxic-ischemic encephalopathy” and a note text search for “therapeutic hypothermia” and related terms such as “hypothermia protocol” and “cooling protocol.” Only patients with CEEG data during the first 5 days of life were included. The obtained patient list was cross validated against an independently curated list of individuals with HIE kept by our Critical Care EEG Service, with manual chart review of selected patients to validate the EMR algorithm.

### Data processing

The diagnostic codes, demographic data (age, sex, race, and ethnicity), and EEG template CDEs were extracted from the EMR and exported using Clarity, a SQL database, and analyzed using the R statistical framework. From these raw data, features from the first day of EEG were selected and mapped onto binary outcomes representing “normal” and “abnormal.” For example, the continuity feature included the levels “low voltage suppressed”, “excessive discontinuity”, “burst suppression”, “normal discontinuity”, and “normal continuity.” “Normal continuity” and “normal discontinuity” were both mapped to “normal,” while the other selections were mapped to “abnormal.” This process was completed for all features and EEG reports. The outcome variable was defined as seizures on subsequent recording days, occurring on day two or later. Patients were considered to have subsequent seizures if there was at least one seizure recorded after the first day. Predictive variables (sex, race, ethnicity, seizures, continuity, variability, reactivity, voltage, graphoelements, epileptiform transients, impression) and outcome measures for each individual were assessed. If neonates had two separate EEG recording periods, separated by at least 24 hours off EEG, then these were treated as two separate sessions. Patients with missing data for any of the key variables were not included in the models.

### Correlation and clustering analyses

Cross-correlation plots were created for all EEG features using the native functions and the *corrplot* library in RStudio. Pearson cross-correlation values were calculated. Features and patients were clustered using the *ComplexHeatmap* package,^26^ separating patients with and without any seizures and using complete linkage clustering and a Euclidean distance measure.

### Model building

Seizure prediction models were developed and tested using logistic regression, decision tree, and random forest analyses (**Table S1**). Given the relative rarity of our outcome of interest (seizure occurrence after day 1), we tuned many of our models to be more sensitive to patients with seizures by training our models with more weight placed on patients who went on to have seizures. We produced logistic regression models with no weights, balanced weights, and by weighting the “subsequent seizure” class three and five times as highly as the “no subsequent seizure” class which was weighted at one half. We produced random forest models with no weights, balanced weights, and by weighting the “subsequent seizure” class between one and a half and 15 times as highly as the “no subsequent seizure” class which was weighted at one half. Logistic regression models were built using the *Caret* package in RStudio. Random forest models were built using the *randomForest* and *ranger* packages as well as the *H2O platform* (H2O.ai, R Interface for H2O, R package version 3.10.0.8. https://github.com/h2oai/h2o-3. Oct. 2016). When evaluating the performance of our classifiers, we used accuracy, precision, recall, F1, area under the curve (AUC), area under the precision-recall curve (AUCPR) and Cohen’s kappa scores. Given our imbalanced data set, we sought to maximize F1 and AUCPR over AUC and accuracy.^27^ Variable importance, which displays the relative influence of each feature, was determined within our H2O models through feature inclusion in a split as well as the squared error reduction at each split in which that feature was included. The larger the reduction in squared error between each node and its children, the higher the variable importance.

## Results

### Standardized EEG reports result in consistent documentation of ACNS-recommended features

Between January 2018 and February 2022, the CDE-based novel documentation system was used to report >42,000 EEGs (**Figure 1A-C**), including 1117 neonates in the intensive care unit. For neonates, we defined key variables as presence or absence of seizures, continuity, variability, reactivity, voltage, graphoelements, epileptiform transients, and impression. Overall, these key EEG features were reported for >95% of reports on the first day of CEEG (**Table 2** and **Figure 2C**).

**Figure 1:**
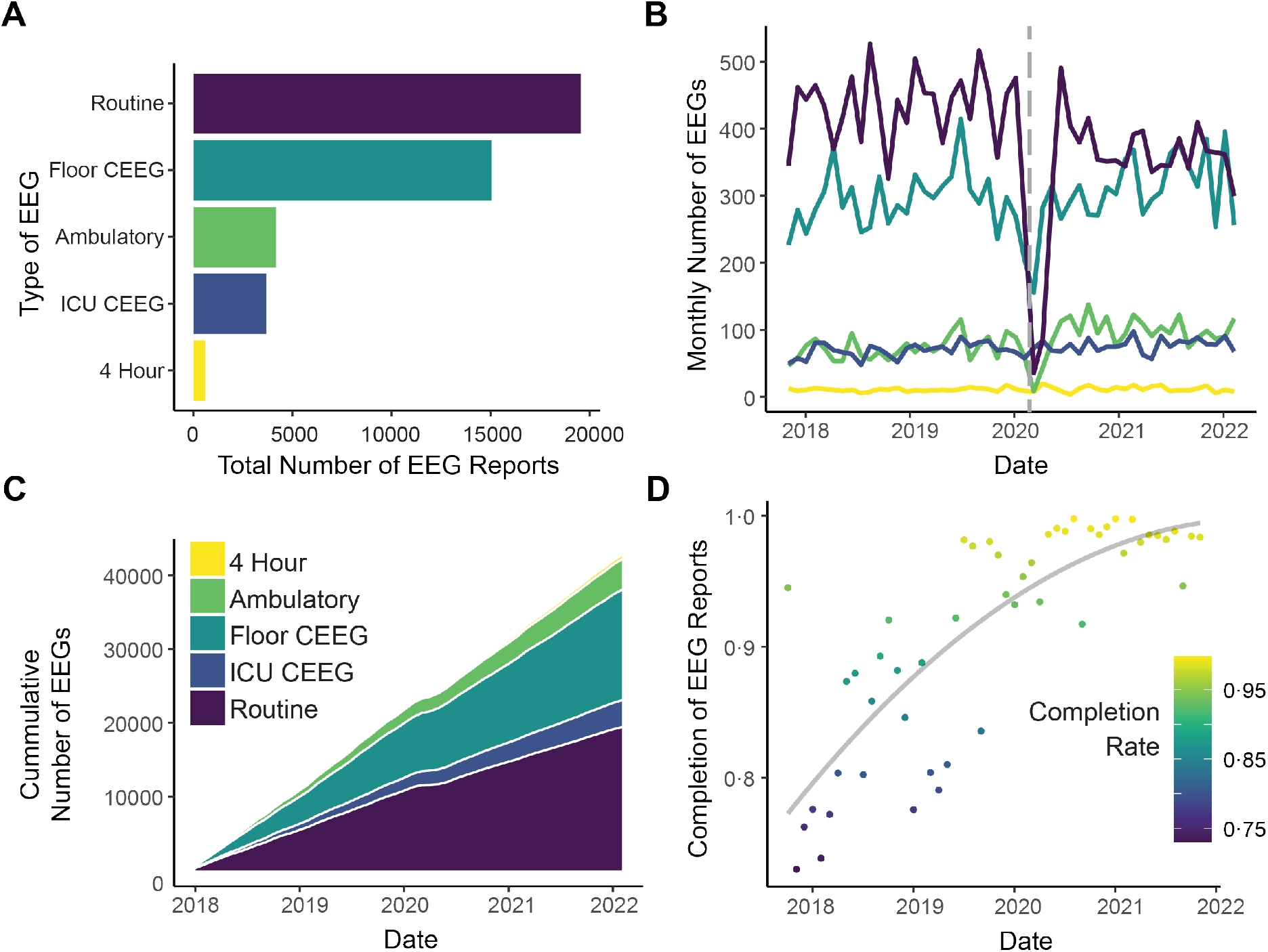
EEG Data Accrual & Completion. (**A**) >42,000 EEGs have been reported using the templated system, most of these being routine (<1 hour) EEGs or hospital-based long-term monitoring (LTM). (**B**) Monthly numbers of EEGs reported using the novel system have remained roughly stable, except during the COVID-19 pandemic (dashed vertical line). (**C**) Cumulative numbers of EEGs continue to grow overtime for all categories. (**D**) Completion index for neonatal CEEGs, defined as proportion of key features described, has improved overtime from <80% when templates were first instituted to >95% currently.

**Figure 2:**
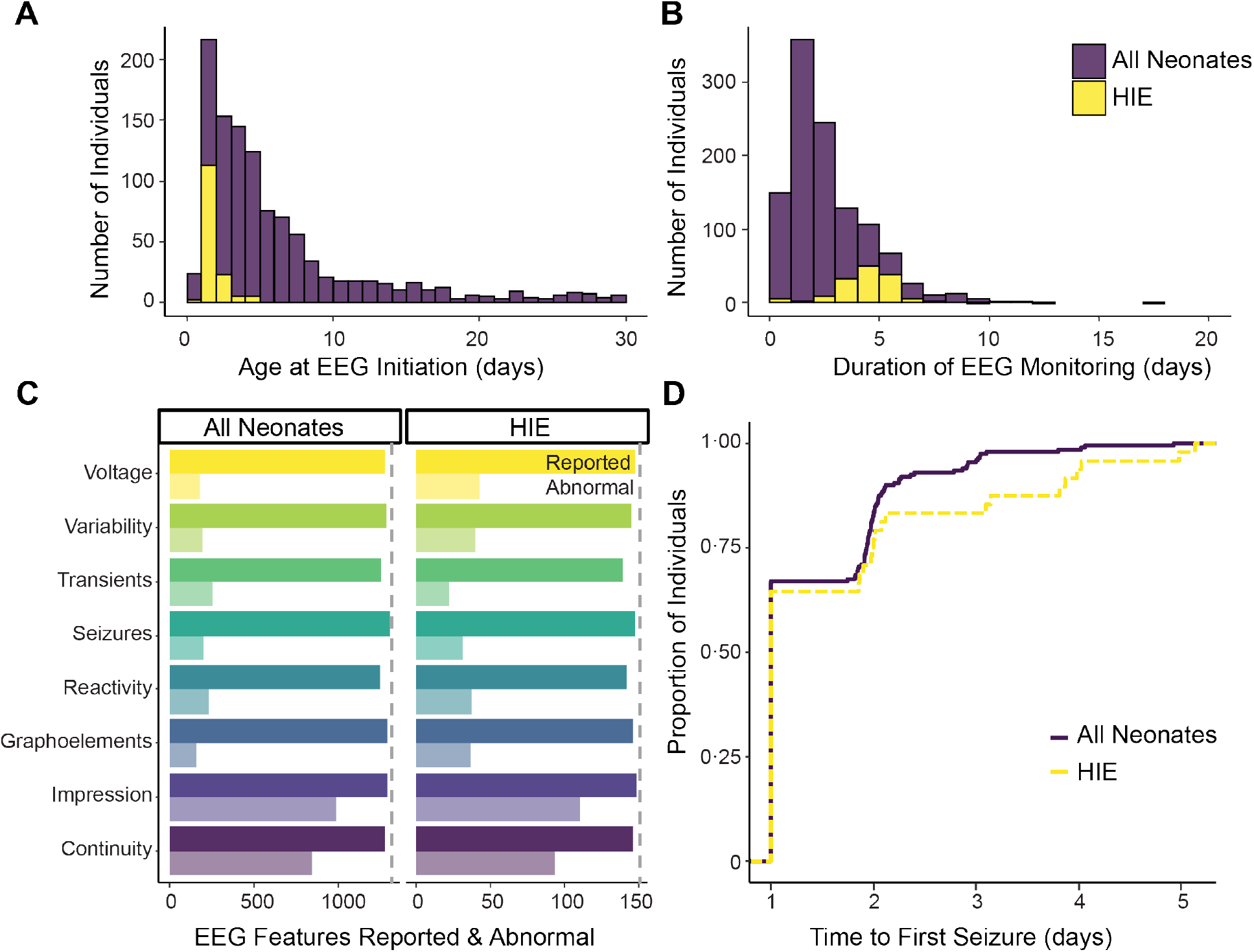
Population characteristics and time to seizure. Distributions of age at EEG initiation (**A**) and duration of EEG monitoring (**B**) for both the overall neonatal population (purple) and the subset of neonates with HIE (yellow). Neonates with HIE had CEEG initiated at younger ages and underwent longer duration CEEG than the overall cohort (see main text for details). (**C**) All key features were reliably reported for both the overall neonatal and HIE cohorts (solid bars). Frequencies of abnormalities varied slightly between the groups (translucent bars). Among the neonates who ultimately had at least one seizure, the duration from CEEG initiation to the first seizure was similar between the two cohorts, with most patients seizing within the first 2 days of monitoring (**D**).

In addition to an overall high completion rate, the completeness of EEG reports improved over time (**Figure 1D**). Neonatal EEGs reported in 2018, during the first 6 months of EMR template use, had an average completion rate of 80% while EEGs reported in 2021-2022, during the most recent 6 months of template use, had completion rates of 98% (p < 0·001, Wilcoxon Rank Sum Test). Therefore, the implementation of EEG templates provided complete data which can be used to build prediction models to inform clinical care.

### Neonates with HIE and therapeutic hypothermia can be reliably identified from medical records

The cohort of neonates who underwent CEEG during the first 30 days of life included 1117 individuals (2/2018 – 2/2022). Demographic features are presented in **Table 1**. The median age of CEEG initiation was 4 days of life (range 0-30 days, **Figure 2A**). The median duration of CEEG was 3 days (range 1-18 days, **Figure 2B**).

**Table 1:**
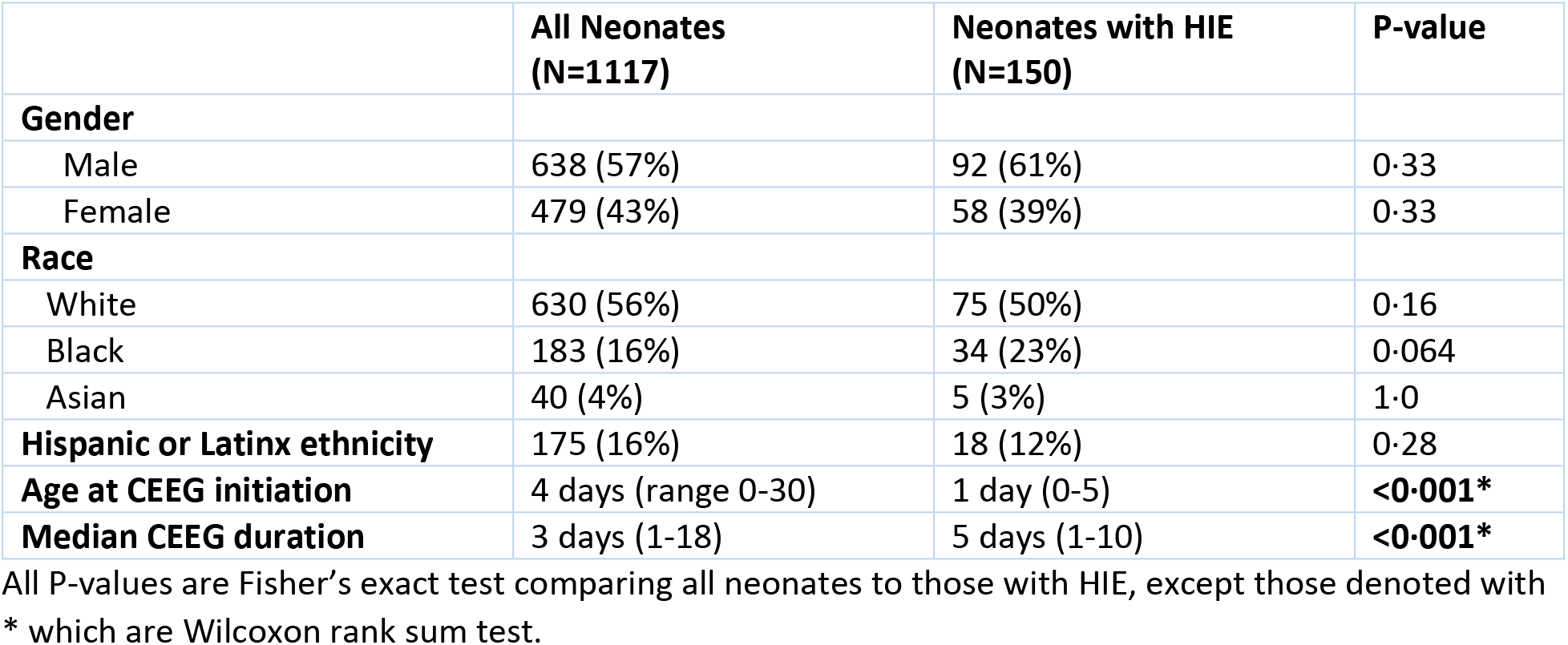
Demographics

Among the 1117 neonates, 150 (13·4%) had HIE managed with therapeutic hypothermia, and these were included in the HIE group for the seizure prediction analyses. Our EMR-based algorithm identified 82/86 (95%) of the individuals from an independent clinical list and also identified 68 additional individuals who met criteria for inclusion. The individuals not captured by this algorithm but identified on the clinical list were patients transferred from outside hospitals (n=2) or patients in whom the diagnosis of either HIE or hypothermia was incorrectly omitted from the diagnosis codes (n=2). Individuals incorrectly selected by our algorithm, including those with initial EEGs performed at outside hospitals or who did not actually undergo therapeutic hypothermia, were manually removed (total n=70). Demographics of the final HIE cohort of 150 individuals are presented in **Table 1** and were not statistically different from those of the broader neonatal cohort, except for age and CEEG duration. The median age at CEEG initiation was 1 day of life (range 0-5 days, **Figure 2A**), and neonates with HIE were significantly younger than the overall cohort (p<0·001, Wilcoxon rank sum test). The median duration of CEEG was 5 days (range 1 – 10 days, **Figure 2B**), and neonates with HIE underwent CEEG for significantly longer than the overall cohort (p<0·001, Wilcoxon rank sum test).

### Neonates with HIE formed a distinct subgroup

We compared the frequency of EEG feature abnormalities between the overall neonatal cohort and the subgroup with HIE. On the first day of recording, individuals with HIE had more background abnormalities (voltage p<0·001, variability p<0·001, reactivity p=0·032, and graphoelements p<0·001) and trended to have more seizures (p=0·097, see **Table 2** and **Figure 2C** for details). Over the course of CEEG, neonates with HIE were also more likely to have seizures, with 33% (n=49) having at least one seizure across all days of recording compared to 26% (n=342) of the overall population, although this difference did not reach statistical significance (p= 0·06, Fisher’s Exact Test). The time to first seizure from CEEG initiation was similar between overall cohort and HIE group (**Figure 2D**).

**Table 2:** Day 1 EEG features

| EEG feature | All Neonates |  | Neonates with HIE |  | P-value* |
| --- | --- | --- | --- | --- | --- |
|  | Number reported | Frequency of abnormalities | Number reported | Frequency of abnormalities |  |
| Continuity | 1283/1313 (97.7%) | 847/1283 (66%) | 146/150 (97.3%) | 93/146 (63.7%) | 0.58 |
| Voltage | 1278/1313 (97.3%) | 180/1278 (14.1%) | 147/150 (98.0%) | 42/147 (28.6%) | **<0.001** |
| Variability | 1289/1313 (98.2%) | 193/1289 (15.0%) | 145/150 (96.7%) | 40/145 (27.6%) | **<0.001** |
| Reactivity | 1247/1313 (95.0%) | 231/1247 (18.5%) | 141/150 (94.0%) | 37/141 (26.2%) | **0.032** |
| Graphoelements | 1293/1313 (98.5%) | 159/1293 (12.3%) | 146/150 (97.3%) | 36/146 (24.7%) | **<0.001** |
| Transients | 1256/1313 (95.7%) | 256/1256 (20.4%) | 139/150 (92.7%) | 22/139 (15.8%) | 0.22 |
| EEG Seizures | 1307/1313 (99.5%) | 203/1307 (15.5%) | 147/150 (98.0%) | 31/147 (21.1%) | 0.097 |
| EEG Impression | 1292/1313 (98.4%) | 989/1292 (76.5%) | 148/150 (98.7%) | 110/148 (74.3%) | 0.54 |

Number reported represents the number of first day EEG reports commenting on the feature of interest. Frequency of abnormalities is the number of first day reports specifying that the feature of interest was abnormal, out of all reports describing the feature. \*Fisher's exact test comparing frequency of abnormalities in all neonates to those with HIE.

### Multiple EEG features in neonates are correlated, but not predictive of future seizures

Several EEG features were correlated for the full cohort and the subgroup with HIE, including overall impression and continuity (0·72 for all, 0.78 for HIE), variability and reactivity (0·44 for all, 0.66 for HIE), voltage and variability (0·39 for all, 0·68 for HIE), and voltage and reactivity (0·43 for all, 0·61 for HIE, **Figure 3A, C**, all comparisons p<0·001). Furthermore, while there were overall differences in abnormal feature representation between the “seizure” and “no seizure” groups, there was a large degree of heterogeneity, and the presence or absence of seizures could not be predicted by feature clustering alone (**Figure 3B, D**). In summary, in addition to being sparsely represented across the cohort, many EEG features were correlated and easily identifiable patterns in the data were not predictive of future seizures. This suggests that the overall structure of the set of reported EEG features is not intuitive and requires more sophisticated modelling approaches to predict future seizures.

**Figure 3:**
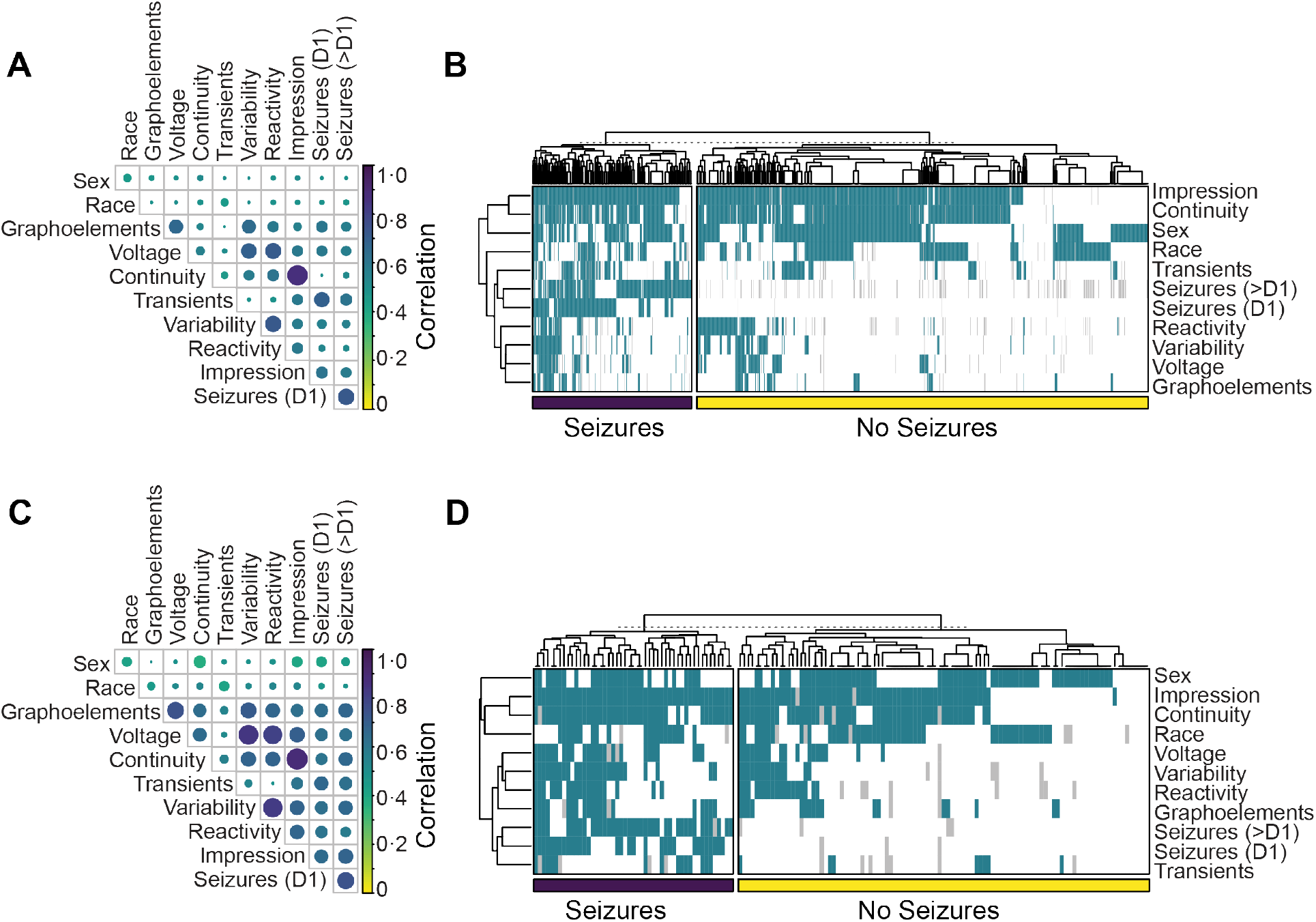
Feature Correlation & Clustering. Cross-correlation coefficients for all EEG features are shown for the entire neonatal cohort (**A**) and the subset with HIE (**C**). Features with the strongest correlations include overall impression and continuity (0.72 for all, 0.78 for HIE), variability and reactivity (0.44 for all, 0.66 for HIE), voltage and variability (0.39 for all, 0.68 for HIE), and voltage and reactivity (0.43 for all, 0.61 for HIE). Clustering of individuals based on EEG feature representation did not clearly segment patients based on the presence or absence of seizures, for the overall cohort (**B**) or the subgroup with HIE (**D**). “Seizures (D1)” are seizures occurring on the first day of EEG recording, while “seizures (>D1)” represent all subsequent seizures.

### Seizure prediction models can identify sub-groups of patients at low or high risk of seizure

A variety of frameworks can be used for prediction models, and we sought to identify the benefits and weaknesses of various methods to optimize the prediction of future seizures. Our primary aim was to build a seizure prediction model to determine which neonates, and particularly which neonates with HIE, would ultimately have seizures, based on EEG features during the first 24 hours of CEEG. We compared (1) logistic regression models, (2) decision trees, and (3) random forest algorithms, representing a shift from standard epidemiological methods to machine learning methods.

#### Logistic regression

Models built with logistic regression predicted subsequent seizures with up to 84% accuracy (0·54 AUCPR) in the full neonatal cohort and 77% accuracy (0·57 AUCPR) in the neonates with HIE (see **Figure 4 & 5** for detailed performance metrics). The highest performing logistic regression model had balanced classes.

**Figure 4:**
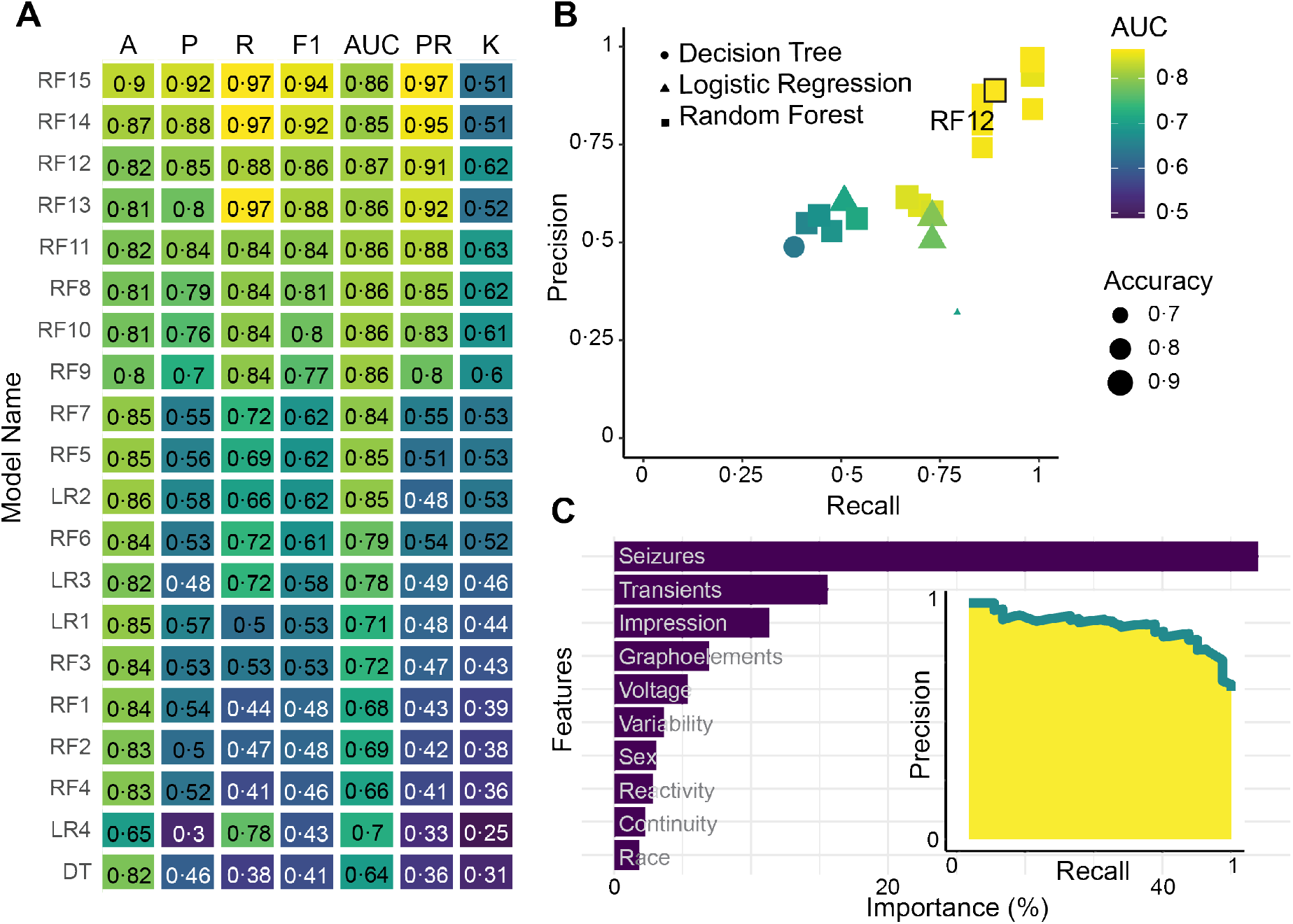
Model Performance for All Neonates. (**A**) Performance values are displayed for the logistic regression (LR, N=4), decision tree (DT, N=1) and random forest (RF, N=15) models tested on the entire neonatal cohort. See Table S1 for model descriptions. Accuracy (A), precision (P), recall (R), F1, AUC, AUCPR (PR), and Cohen’s Kappa (K) scores are provided for each model. Color represents performance. (**B**) The precision of each model plotted over recall for all 20 models, coded by type, AUC and accuracy (see legend). (**C**) The relative importance of each feature in the model, as well as the precision-recall curve are shown for model RF12.

**Figure 5:**
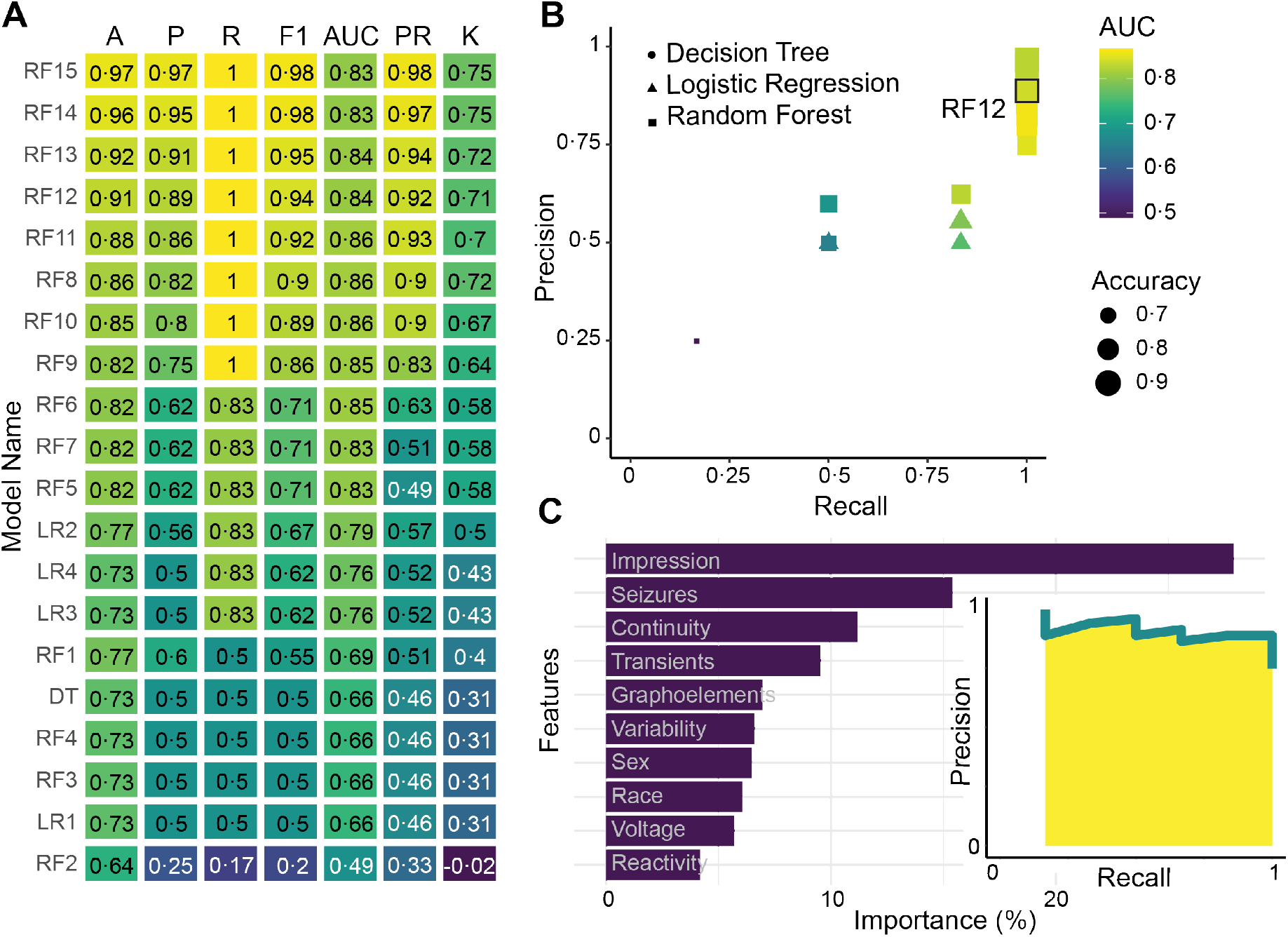
Model Performance for Neonates with HIE. (**A**) Performance values are displayed for the logistic regression (LR, N=4), decision tree (DT, N=1) and random forest (RF, N=15) models tested on the HIE cohort. See Table S1 for model descriptions. Accuracy (A), precision (P), recall (R), F1, AUC, AUCPR (PR), and Cohen’s Kappa (K) scores are provided for each model. Color represents performance. (**B**) The precision of each model plotted over recall for all 20 models, coded by type, AUC and accuracy (see legend). (**C**) The relative importance of each feature in the model, as well as the precision-recall curve are shown for model RF12.

#### Decision trees

Decision trees, which are supervised machine learning algorithms useful for solving classification problems with categorical variables, are simple, intuitive, fast, and not influenced by outliers or missing data.^28^ An example decision tree with 92% classification accuracy is shown in **Supplemental Figure 3**. One preferred feature of these models is the ability to identify patients at low and high risk of seizures by examining the leaves of the tree. Despite these advantages, a basic decision tree model did not outperform logistic regression, with accuracies of 82% (73% for those with HIE) and AUCPR of 0·36 (0·46 for those with HIE, **Figure 4 & 5**). Thus, while decision trees are intuitive and could identify groups of patients at high and low risk of subsequent seizures more readily than logistic regression models, they still had suboptimal performance overall.

#### Random Forests

Random forest models are built using ensembles of many decision trees. The assumption underlying these models is that large numbers of uncorrelated trees operating as a group will outperform any constituent model. This prevents overfitting, which can occur with decision trees, and improves the stability of the final model. Untuned random forest models performed our classification task with accuracies similar to regression models. However, tuned models that more heavily weighted the subsequent seizure class help the model identify these patients with much greater success, resulting in recall of up to 97% in the overall neonatal cohort and >99% in the subgroup with HIE. AUCPR values were also excellent, up to 0·97 for all neonates and 0·98 for neonates with HIE. This class weighting came at a cost of lower prediction accuracy for the null class (those that do not have seizures); however, the accuracies of these models were still up to 90% for the overall cohort and 97% for the subgroup with HIE (**Figure 4 & 5**). Additionally, for clinical applications, this represents a strength of this model, as it can detect which neonates will have seizures with very high sensitivity, while still identifying a sufficiently large proportion of neonates at low risk of seizure. Therefore, random forest models had higher overall performance metrics than regression and decision tree models and identified patients who subsequently had seizures with recall approaching 100%.

## Discussion

In our study, we aimed to assess whether a standardized EEG reporting template facilitates neonatal seizure prediction models to guide clinical care. Using the entirety of neonatal EEG reports collected over a four-year period at a large tertiary pediatric center, we find that neonatal seizures on subsequent days after initial recording are highly predictable. Our results provide the basis for rational continuous EEG (CEEG) use, particularly in limited settings. For example, our findings suggest that a center without the ability to monitor HIE patients with CEEG through rewarming (4-5 days) could perform 24 hours of CEEG and be assured that subsequent seizures would not be missed in neonates identified as low risk.

Our study has three main findings. First, we demonstrate that using a standardized EEG reporting template in the electronic medical record (EMR) dramatically improves compliance with recommended terminology,^24^ increasing from 80% to near complete compliance of 98%, with a gradual improvement over time. This result is surprising and speaks to a dynamic interaction between tool implementation and provider uptake: even though compliant use of the new format was neither monitored nor reinforced, EEG readers slowly gravitated towards a more complete reporting format. This example demonstrates how implementation of novel technologies can provide a subtle transformative effect in areas of healthcare where exact reporting is critical.

Second, as the main clinical finding of our study, we demonstrated that clinical data from ongoing care can predict neonatal seizures on subsequent days with high accuracy. This finding speaks to the overall power of learning health system approaches in child neurology, which lags behind other fields in medicine.^29^ With only limited existing knowledge about predictive factors, the considerable data from routine care can be used to make reliable predictions. Even in specific clinical scenarios such as neonatal CEEG recordings, standardized reporting templates can generate sufficient data within a four-year period to allow for adequate accuracy to be incorporated into clinical care. Of note, our predictions were made based on standardized EEG reports rather than raw EEG data. The strong predictive power of this data supports the value of such information, which might be considered only indirect at first. However, each EEG report represents the expert assessment of a certified EEG reader, thereby allowing us to utilize a data resource that is highly informative and valuable.

Third, we found that even for standardized datasets such as EEG reports, the use of machine learning methods such as random forests surpasses the use of conventional statistical approaches such as logistic regression. This finding and the lack of predictive power of single EEG features can be attributed to the complexity inherent in EEG reports. Many EEG features are highly correlated, which represents an ongoing challenge for traditional statistical approaches. Tools such as random forests are less susceptible to such confounding structures and can therefore generate more accurate predictions.

Inferences about underlying causes and biology are limited in machine learning approaches in contrast to single feature predictions or logistic regression. For example, with decision trees, we frequently observed trees providing comparable classifications with completely different decision hierarchies. This indicates that model performance may be considered separate from conclusions about underlying mechanisms, which might be better addressed by other study designs. In contrast to simple decision trees that can be visualized and followed to guide clinical practice, random forest models do not have a simple or intuitive visual representation, i.e., the term “model” does not refer to a single visual tree that can be provided as a flowchart. We therefore created an online calculator (available at http://neopredict.helbiglab.io) that generates predictions based on the most recent analysis, which is updated on a quarterly basis with the most recent data in incremental intervals, following the paradigm of a learning health system.^29^

In addition to seizure prediction in all neonatal recordings, we specifically assessed prediction models in neonates with hypoxic-ischemic brain injury. Our results show that even for smaller cohorts, data from ongoing care can correctly identify all neonates with seizures on subsequent days without negatively impacting precision. This finding suggests that predictions in smaller sub-cohorts is feasible alongside the analysis of the larger cohort. In the future, the efficacy of seizure prediction may be improved further by considering additional data already existing in the EMR beyond demographic features such as sex and race included in our current models. For instance, clinical variables in combination with EEG features have previously resulted in improved model performance. For a seizure prediction model built using Cox’s proportional hazard regression, the AUC improved from 76.4% to 83% when clinical features, including gestational age, EEG indication and etiology/therapies, were added to EEG-based prediction alone.^16^ Features such as gestational age, Apgar scores, diagnosis and phenotypic features such human phenotype ontology (HPO) codes,^30^ physical exam findings, medication administration, and laboratory data are increasingly standardized within the EMR. As these data accrue over time, we anticipate that it will be feasible and beneficial to incorporate them into our predictive models.

In summary, we have built the first high-performing neonatal seizure prediction models using automatically extracted EEG documentation from the EMR, achieving accuracies of >90%. We demonstrate that the use of routine clinical care data alone without prior assumptions is sufficient for meaningful seizure predictions in at-risk neonates. Incorporating accurate seizure prediction into real-time clinical care can improve the quality and efficiency of care for critically ill neonates.

## Data Availability

Data in a de-identified format will be made available by request to the corresponding author.

http://neopredict.helbiglab.io

## Data Sharing

Data in a de-identified format will be made available by request to the corresponding author.

## Author Contributions

<u>Jillian L. McKee</u> - Conceptualization, Data Curation, Investigation, Methodology, Formal Analysis, Validation, Visualization, Writing – original draft, Writing – review & editing, Software

<u>Michael C. Kaufman</u> – Data Curation, Investigation, Methodology, Formal Analysis, Validation, Visualization, Writing – review & editing, Software

<u>Alexander K. Gonzalez</u> - Data Curation, Methodology, Software

<u>Mark P. Fitzgerald</u> - Conceptualization, Writing – review & editing, Supervision

<u>Shavonne L. Massey</u> - Data Curation

<u>France Fung</u> - Data Curation, Writing – review & editing

<u>Sudha K. Kessler</u> - Data Curation

<u>Stephanie Witzman</u> - Data Curation, Software

<u>Nicholas S. Abend</u> - Conceptualization, Writing – review & editing, Supervision

<u>Ingo Helbig</u> - Conceptualization, Methodology, Writing – review & editing, Supervision, Funding acquisition, Resources

