## Supplemental Information for "Seizure prediction in 1117 neonates leveraging EMR-embedded standardized EEG reporting"

### McKee 2022 Supplemental Information

**Table of Contents**


Figure S1 Neonatal EEG Template……………………………………………………………………….Page 1
Figure S2: Kaplan-Myer Survival Analysis……………………………………………………………….Page 2
Figure S3: Example decision tree for HIE patients……………………………………………………….Page 3
Table S1: Model Methods……………………………………………………………...…………………Page 4


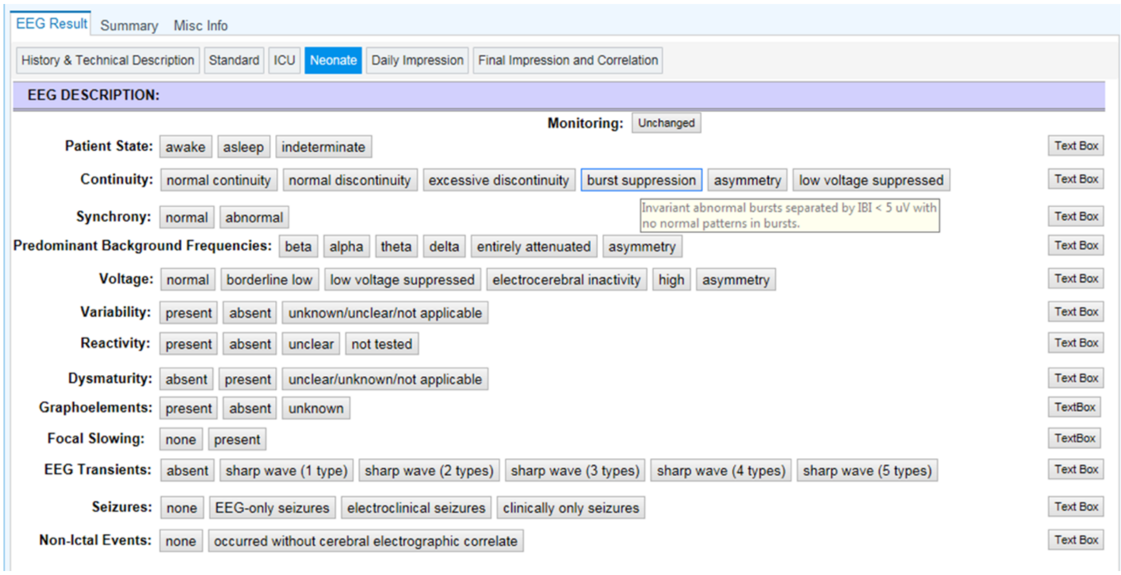


**Figure S1: Neonatal EEG Template.** EEG variables are derived from American Clinical Neurophysiology Society standardized EEG terminology, and hover boxes define terms to users.

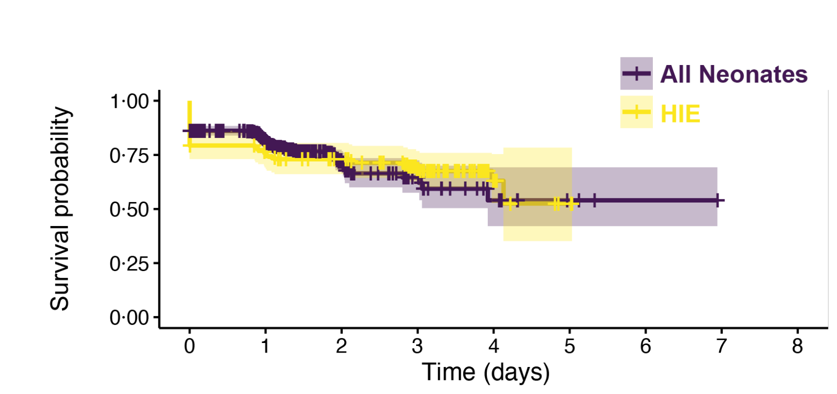


**Number at risk**

| | | | | | | | | | |
|---|---|---|---|---|---|---|---|---|---|
| All Neonates | 967 | 488 | 67 | 27 | 10 | 3 | 1 | 0 | 0 |
| HIE | 150 | 107 | 94 | 65 | 11 | 2 | 0 | 0 | 0 |

**Cumulative number of events**

| | | | | | | | | | |
|---|---|---|---|---|---|---|---|---|---|
| All Neonates | 133 | 165 | 192 | 197 | 200 | 200 | 200 | 200 | 200 |
| HIE | 31 | 36 | 40 | 45 | 47 | 48 | 48 | 48 | 48 |

**Figure S2: Kaplan-Myer Survival Analysis.** Proportion of individuals with seizure-free survival is displayed for both the entire cohort (yellow) and those with HIE (blue). Individuals are censored when monitoring is discontinued (vertical marks).

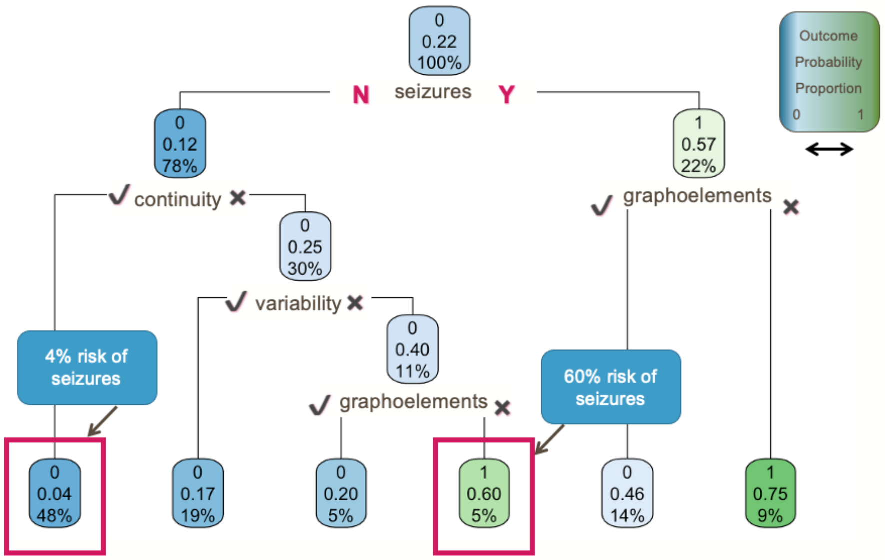


**Figure S3: Example decision tree for HIE patients.** The model is initiated with all patients and 22% risk of seizures (top). This tree then divides patients based on the presence or absence of seizures on day 1. For example, among patients who did not have seizures on day 1 (on the left), only had a 12% risk of seizures on subsequent days. Next, the model recursively splits the population based on the other features to create the branches, until it reaches a terminal leaf that is either homogenous in outcome or too small to split further. The highlighted group on the left accounts for 48% of the population, but only has a 4% risk of future seizures. However, the highlighted group on the right, while only representing 5% of the population has a 60% chance of future seizures.

#### Table S1: Model Methods

| Key | Model Name | Model Type | Description |
|---|---|---|---|
| LR1 | log_regress_caret | Logistic Regression | Default logistic regression model with k-fold cross-validation (k=10) using the Caret package in R. |
| DT | regresstree_caret | Decision Tree | Default decision tree model with cross-validation (k=10) using the Caret package in R. |
| RF1 | random_model1 | Random Forest | Default random forest model using the randomForest package in R. |
| RF2 | random_model_mtry | Random Forest | Random Forest model, optimized for minimal OOB Error using stepwise tuning of mtry. Optimal mtry = 7. |
| RF3 | random_model_opt | Random Forest | Random Forest model, optimized for minimal error rate. Parameters tested included mtry (range 1-10, by increments of 1), minimal node size (3-9, by increments of 2) and number of trees (250-500, by increments of 50). Optimal mtry = 4, optimal node size = 9, optimal number of trees = 500. |
| RF4 | range_1 | Random Forest | Random forest model using the Ranger package in R optimized for OOB error rate. Parameters tested included mtry (range 1-10, by increments of 1), minimal node size (3-9, by increments of 2), sample size (0·55, 0·632, 0·7, 0·8), and number of trees (250-500, by increments of 50). Optimal mtry = 8, optimal node size = 3, optimal sample size = 0·632, optimal number of trees = 250. |
| RF5 | h2o_1 | Random Forest | Distributed Random Forest model using the H2O package in R. Parameters tested included mtry (range 1-10, by increments of 1),sample size (0·55, 0·632, 0·70, 0·80), and number of trees (200-500, by increments of 100). The model was optimized towards maximum AUCPR. Optimal mtry = 2, optimal sample size = 0·55, optimal number of trees = 400. |
| RF6 | h2o_balanced | Random Forest | Distributed Random Forest model using the H2O package in R. Parameters tested included mtry (range 1-10, by increments of 1), sample size (0·55, 0·632, 0·70, 0·80), and number of trees (200-500, by increments of 100). In order to create balance, the model was stratified and the class balance default parameter was activated. The model was optimized towards maximum AUCPR. Optimal mtry = 2, optimal sample size = 0.80, optimal number of trees = 200. Cross-validation (k=10) was also implemented within the model. |
| RF7 | h2o_custom_bal | Random Forest | Distributed Random Forest model using the H2O package in R. Parameters tested included mtry (range 1-10, by increments of 1), sample size (0·55, 0·632, 0·70, 0·80), and number of trees (200-500, by increments of 100). In order to create balance, the model was stratified and the class balance parameter was activated with “no subsequent seizures” undersampled at a rate of 0·5 and “subsequent seizures” sampled at a rate of 0·9. The model was optimized towards maximum AUCPR. Optimal mtry = 1, optimal sample size = 0·70, optimal number of trees = 400. Cross-validation (k=10) was also implemented within the model. |
| RF8 | h2o_weighted_0.6068152_2.840491 | Random Forest | Distributed Random Forest model using the H2O package in R. Parameters tested included mtry (range 1-10, by increments of 1), sample size (0·55, 0·632, 0·70, 0·80), and number of trees (200-500, by increments of 100). In order to create balance, the model was stratified and weighted in order to proportionally distribute points to “non-subsequent seizure” (0·61) and “subsequent seizure” (2·84) instances. The model was optimized towards maximum AUCPR. Optimal mtry = 1, optimal sample size = 0·55, optimal number of trees = 200. Cross-validation (k=10) was also implemented within the model. |
| RF9 | h2o_weighted_0.5_1.5 | Random Forest | Same as above model, aside from weighted metrics for “non-subsequent seizure” (0·5) and “subsequent seizure” (1·5). Optimal mtry = 1, optimal sample size = 0·55, optimal number of trees = 200. |
| RF10 | h2o_weighted_0.5_2 | Random Forest | Same as above model, except weights for “non-subsequent seizure” (0·5) and “subsequent seizure” (2·0). Optimal mtry = 1, optimal sample size = 0·55, optimal number of trees = 200. |
| RF11 | h2o_weighted_0.5_3 | Random Forest | Same as above model, except weights for “non-subsequent seizure” (0·5) and “subsequent seizure” (3·0). Optimal mtry = 1, optimal sample size = 0·55, optimal number of trees = 200. |
| RF12 | h2o_weighted_0.5_4 | Random Forest | Same as above model, except weights for “non-subsequent seizure” (0·5) and “subsequent seizure” (4·0). Optimal mtry = 1, optimal sample size = 0·55, optimal number of trees = 200. |
| RF13 | h2o_weighted_0.5_5 | Random Forest | Same as above model, except weights for “non-subsequent seizure” (0·5) and “subsequent seizure” (5·0). Optimal mtry = 1, optimal sample size = 0·55, optimal number of trees = 200. |

| | | | |
|---|---|---|---|
| RF14 | h2o_weighted_0.5_10 | Random Forest | Same as above model, except weights for “non-subsequent seizure” (0·5) and “subsequent seizure” (10). Optimal mtry = 1, optimal sample size = 0·55, optimal number of trees = 300. |
| RF15 | h2o_weighted_0.5_15 | Random Forest | Same as above model, except weights for “non-subsequent seizure” (0·5) and “subsequent seizure” (15). Optimal mtry = 1, optimal sample size = NA, optimal number of trees = 200. |
| LR2 | log_regress_caret_wb | Logisitc Regression | Logistic regression model with cross-validation (k=10) using the Caret package in R. Weights were added to proportionally distribute points to “non-subsequent seizure” (0·61) and “subsequent seizure” (2·84). |
| LR3 | log_regress_caret_w3 | Logisitc Regression | Same as above model, aside from weighted metrics for “non-subsequent seizure” (0·5) and “subsequent seizure” (3·0) |
| LR4 | log_regress_caret_w5 | Logistic Regression | Same as above model, aside from weighted metrics for “non-subsequent seizure” (0·5) and “subsequent seizure” (5·0) |
